# Admission systolic blood pressure and obesity correlate with fatal and severe acute COVID-19 in the population of New Orleans, LA

**DOI:** 10.1101/2024.06.27.24309604

**Authors:** Dahlene Fusco, Sharon Liu, Marc Theberge, Anuhya V. Pulapaka, Yitian Zha, William Rittmeyer, Marlowe Maylin, W. Ben Rothwell, Prateek Adhikari, Peter Raynaud, Keith Ferdinand, Arnaud Drouin

**Affiliations:** Tulane University School of Medicine, New Orleans Louisiana; University Medical Center, New Orleans Louisiana; Tulane University, New Orleans Louisiana; Tulane Medical Center, New Orleans Louisiana

**Keywords:** COVID-19, SARS CoV-2, hypertension, obesity, clinical predictor of outcomes, clinical triage, ClinSeqSer acute COVID-19 observational study

## Abstract

**BACKGROUND:** In New Orleans, Louisiana (NOLA), the population’s very high social vulnerability led to the establishment of an early epicenter for severe acute COVID-19. Anticipating future respiratory virus outbreaks, identifying low-cost correlates of outcome relevant to special populations is crucial.

**METHODS:** 89 patients with acute COVID-19, enrolled March to August 2020 in the ClinSeqSer longitudinal observational study.

**RESULTS:** The cohort’s population, ∼70% Black, 53% female and 55% obese, reflects exactly that of greater urban NOLA; In contrast, pre-COVID hypertension (HTN) is 83% and 1.5 to 2-fold the state’s prevalence (43% among White, 56% among Black residents). Black patients are younger than White (∼50% vs <= 30% in 45-64 years age bracket). Outcomes were 47% severe, including 17% fatal, and 30% non-fatal (high flow or intubated), and identical by race/age. Obesity, BMI, admit systolic blood pressure (SBP), pulse BP, and CRP level, but not race, sex, age, type 2 diabetes, HTN, number or specific anti-HTN drugs, correlated with fatal and severe outcomes. Patients with admission SBP ≥140 mmHg reached severe clinical state sooner than those with lower SBP.

**CONCLUSIONS:** The very high proportion of pre-COVID HTN in this acute COVID cohort correlates with high social vulnerability. Obesity and SBP on admission stand out as risks for fatal and severe outcomes of acute COVID. The findings support further study of acute COVID admit SBP as a potential correlate of outcome, and the potential role for interactions between a single strain of SARS CoV-2 and the renin-angiotensin-aldosterone blood pressure axis.

## BACKGROUND

Evaluation of acute coronavirus infectious disease 2019 (COVID) cohorts from the pre-vaccine era may provide insights into increasingly recognized acute and long-term effects of SARS CoV-2 on patients’ cardiovascular system [1]. The current study presents pre-vaccine COVID data from a region consistently ranked among U.S. worst for cardiovascular outcomes. The first case of COVID was diagnosed in New Orleans, LA (NOLA) March 9^th^, 2020, just after the end of the Mardi Gras festivities where a massive spread of a single strain occurred over a couple of weeks instead of multiple simultaneous strains as observed in other US cities [2, 3]. NOLA was declared an early U.S. epicenter of COVID by the end of March 2020, and by July 2020, 116,280 total cases and 3,835 deaths from COVID were reported [4]. The NOLA population is comprised of ∼60% Black and ∼36% White residents with a very high Social Vulnerability Index, at 0.98 (0 – 1 scale) [5]. In addition, LA ranks among U.S. states with the 3rd highest proportion of population enrolled in Medicaid (41%) and a share of people living in poverty (19%) [6]. The population in LA is cited as one of the least healthy in the U.S. [7] and life expectancy in 2017 was 73 years, ∼3 years lower than U.S. average, and by race/sex strikingly different for White (male 80-85;female > 85), versus Black (male < 70; female 75) residents [8]. LA ranks fifth in the U.S. for death by heart disease [9], with high prevalence of obesity (45-55%) compared to overall U.S. population (∼35%). LA prevalence of essential hypertension (HTN) (Black 56%; White 48%) is also higher than general U.S. prevalence (48%). High rates of cardiovascular (CV) disease observed in LA occur at relatively young age [10, 11], illustrated by the high percentage and young age of limb amputations in patients with type two diabetes mellitus (T2DM) [12]. The Bogalusa Heart Cohort study has documented CV disease in rural LA since the 1970s and determined that Black residents have very high CV risk, with obesity and HTN developing early in childhood [13–15].

The cohort illustrates the CDC statement that “The pandemic has highlighted racial, ethnic, and socio-economic disparities in COVID-19 illnesses, hospitalizations, and deaths” [16] as marked inequity underlined by racial disparities paralleled COVID mortality in LA [17] and in the U.S. by the decrease in life expectancy by ∼2 years from 2018 to 2020 (Black Americans lost 3.25 years while White Americans lost 1.36 years) [18].

Variants of SARS CoV-2 will likely evolve and circulate for months to years to come, underscoring a critical need for simple clinical outcome predictors, immediately available on admission. In pursuit of identifying such predictors, we present data from the ClinSeqSer acute COVID observational study, conducted in two medical centers in NOLA at the beginning of the regional outbreak.

## METHODS (Refer also to Supp. File #1 Methods)

### Study type

ClinSeqSer is an observational, longitudinal, regional study, initiated March 2020, aimed to define correlates of acute COVID outcomes with any of the following: pre-COVID Long Term Conditions (LTC) and medications, presenting vitals, clinical evolution during admission, and clinical laboratory results. Recruitment: Adult patients ≥18 years old were approached based upon positive nasopharyngeal SARS CoV-2 genome PCR detection, followed by informed consent. Enrollment occurred in person during hospital admission.

### Ethics approval

Study was conducted with approval from Tulane University School of Medicine Institutional Review Board (IRB) under protocol “Collection of SARS CoV-2 Serum and Secretions for Countermeasure Development” [IRB 2020-396, approved 3/20/2020]. All procedures were performed in accordance with ethical standards of Tulane IRB and Helsinki Declaration of 1975 (revised 1983).

### Clinical trial registration number

NCT04956445.

### Data source and collection

Enrollment sites included Tulane Medical Center (TMC) and University Medical Center (UMC), both in NOLA. Health insurance coverage, ZIPcode race/sex, clinical data and laboratory test results were collected from electronic health records (eHR) MEDITECH (TMC) and EPIC (UMC) and stored in REDCap. To optimize homogeneity and accuracy of data, extraction was performed with entry verification for ∼20% of cases, randomly selected, verified by independent study team members, and correlation debriefing meetings.

### ZIP code Health insurance, race and sex

Collected in the eHR under section “demographics”. Race is self-reported, Black patients or African American non-Hispanic, White or Caucasian, and Hispanic Latino-American (Latinx). Because of age and comorbidity similarities, we included the two Latinx patients within the group of Black patients.

### Body Mass Index (BMI)

Collected from admission or medical visits within the six months preceding COVID admission, kilogram weight/square meter height (kg/m^2^). Obesity i**s** defined by WHO criteria as BMI ≥30kg/m^2^, super-obese as BMI ≥40kg/m^2^.

### Essential Hypertension (HTN)

Defined as systolic blood pressure (SBP) ≥130 mmHg [19], with ICD10 code for HTN, present prior to COVID diagnosis, reported in pre-COVID clinical notes in two prior visits for more than 80% patients admitted. Medications prescribed for treatment of HTN were also collected from eHR. For patients without prior visits recorded, presence of HTN/anti-HT medications was collected from History and Physical (H&P) on admission.

### Co-morbidity scores

**1-The Charlson comorbidity Index score (CCI),** modified Deyo version, is independent of age, and scores 1-29. **2-The Elixhauser Index score**, van Walraven algorithm, is provided either by range of likelihood for in-hospital death (−19 (less) to 89 (more)), or by percent of possible points (0 to 82.4%). Calculators for both scores are available free online (orthotoolkit.com).

**Clinical scores** were stratified per WHO *Clinical score evaluation* on a seven-point ordinal scale [20]. S**evere and non-severe groups:** patients with clinical scores of 1 to 3 are included in **“fatal & severe”,** hereafter grouped together as **“severe”** unless otherwise specified, and 4 to 7 in **“non-severe”** COVID outcome groups (Supp. Methods file).

### Blood pressure (BP) on admission

Systolic and diastolic BP were collected from the first vitals registered by emergency medical services or in emergency department. To avoid false high readings when regular size arm cuffs are used in obese patients, TMC and UMC adhere to policies requiring use of large arm cuffs. We excluded values when patients were on oxygen supplement or mechanically ventilated prior to transfer from an outside hospital or profoundly hypotensive on admission (SBP ∼90 +/-2 mmHg, MAP <70 mmHg), all confirmed in eHR.

### Statistical analysis

We report categorical variables as percentages, continuous variables as mean/SD if normally distributed or median/IQR if non-normally distributed. Data were analyzed using Chi-squared, Mann Whitney U test or t-test and Z-test methods as appropriate. Statistics applied for normal distribution assessment, average/median/IQR, ROC curve analysis were performed in Excel, for t-test, Mann Whitney, two proportion Z-test calculator using the calculator tools of Statology [21], for relative risk the calculator from MedCalc [www.medcalc.org], and for Kaplan-Meier log-rank test and graphs the Statistics Kingdom website and R and RStudio program [21–23].

## RESULTS

From March to August 2020, of the 89 patients recruited, ∼70% are Black, 54% female, and, on average, 66 years old (**Table 1**). ZIP code and eHR indicate that the cohort’s Black patients live in areas of higher social vulnerability [2, 3] with 30% enrolled in Medicaid vs 21% national average [17]. Enrolled Black male and female patients are younger than White males (∼50% vs ≤30% in 45-64 years age bracket), while most White females are 85 and older (**Table 1**, **Fig.1**). Patients’ outcomes are 17% fatal, 30% severe non-fatal (intubated or high flow oxygen), and 53% non-severe.

**Figure 1.**
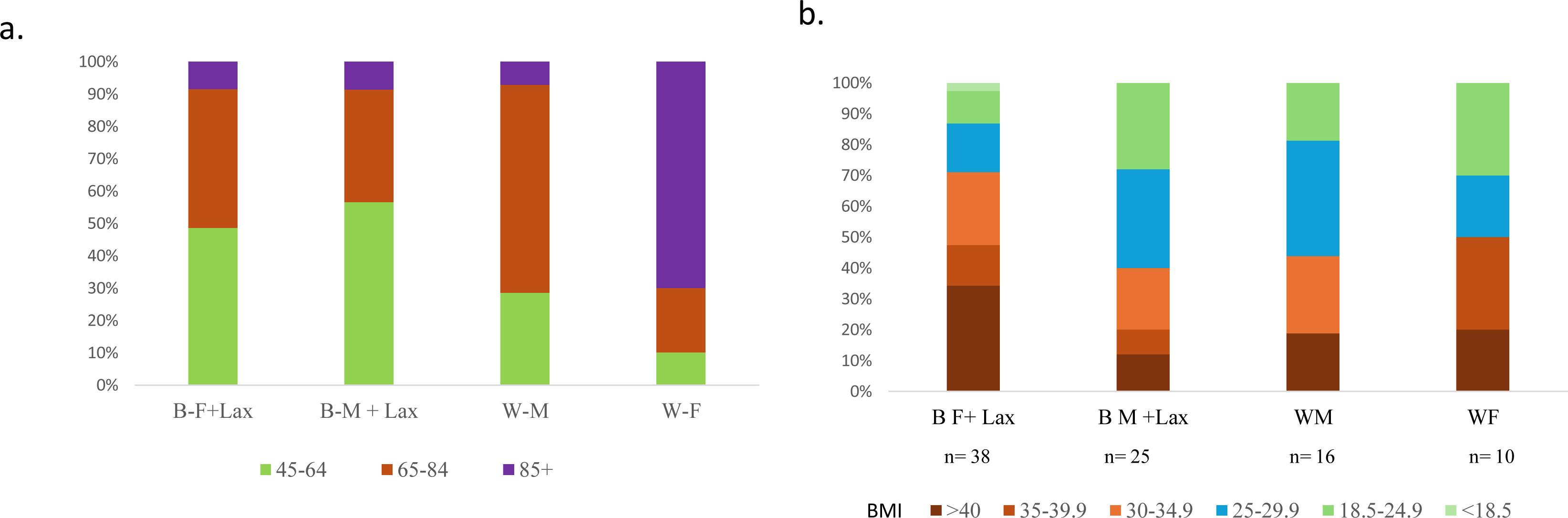
Distribution of age and pre-COVID obesity in the ClinSeqSer Cohort. Race/sex groups (horizontal axis) represent Black (B), Latinx (Lax), and White (W) males (M) and females (F), patients by **a-** age brackets, and **b-** BMI (kg/m^2^) stratification, vertical axis= percent (%). n= number of patients per race and sex group.

**Table 1.**
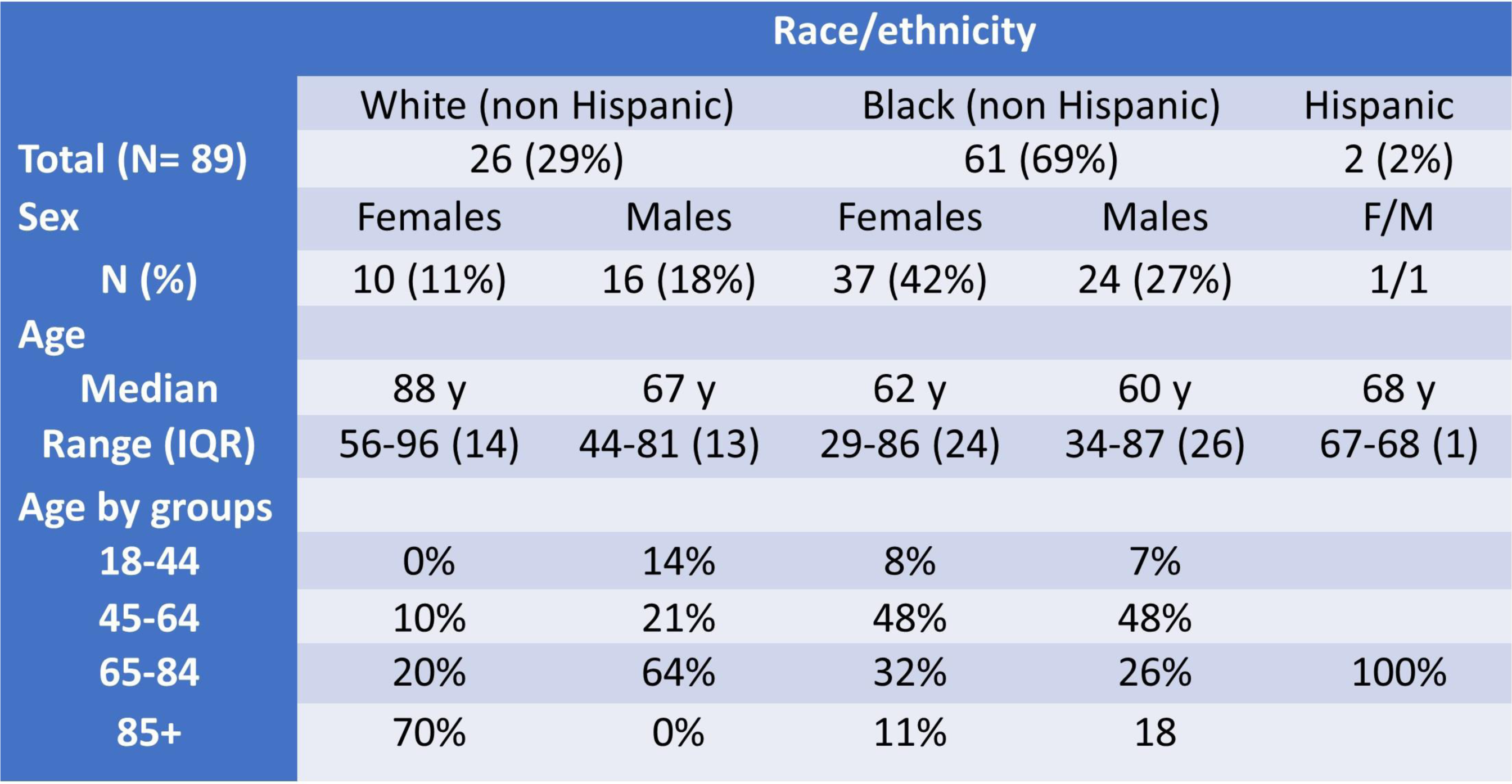
General Demographics of ClinSeqSer cohort, including race, ethnicity, sex, age.

The overall proportion of obese patients is 55%, and the median BMI is 31 kg/m^2^ both significantly higher in Black patients, 63% and 33 kg/m^2^ (**Fig.1**). BMI median values correlate with score of severity (p= 0.002), while proportion of patients with obesity is significantly different in non-severe vs. severe outcomes (**Table 2**) and obesity is a 2.2x relative risk of severe outcome (p= 0.0075, 95% CI 1.2-3.4) (**Table 3**).

**Table 2.** Characteristics of patients with prior to COVID-19 HTN by outcomes. Clinical Scores 1-3 = fatal & severe, CS 4-7= non severe, demographics, associated co-morbidity, HTN on admission and anti-HT therapy in patients with HTN. Statistical tests used: *chi sq ** t test *** z test tow tailed).

|  | Severe COVID | Non-severe COVID | P values |
| --- | --- | --- | --- |
|  | **47% (n= 42)** | **53% (n= 47)** |  |
| F / M ratio | 1.0 | 1.35 | ns** |
| Black+Latinx / White ratio | 2.2 (29 vs 13 ) | 2.6 (34 vs 13) | ns** |
| Age y AVE (SD) | 67 (+/-13) | 71 (+/-15) | ns* |
| BMI (kg/m<sup>2</sup>) median (IQR) | 35 (13) | 27 (10) | p=.002* |
| Obesity BMI ≥30kg/m<sup>2</sup> | 71% (30/42) | 40% (19/47) | p=.0046** |
| Type 2 diabetes | 31% (13/42) | (17/47) 36% | ns** |
| Charlson's score | 3.7 | 3.6 | ns** |
| Elixhauser score | 81% | 77% | ns** |
| Prior COVID HTN | 90% (38/42) | 76% (36/47) | ns (p=.08)*** |
| HT therapy, no drug | 16% (6/38) | 16% (7/36) | ns*** |
| ≥ 1 anti-HT drug | 84% (32/38) | 70% (25/36) | ns*** |
| One anti-HT drug | 50% (19/38) | 48% (15/36) | ns*** |
| ≥2 anti-HT drugs | 34% (13/38) | 39% (10/36) | ns*** |
| **On admission systolic and diastolic blood pressure average (med. IQR)** |  |  |  |
| Systolic (mmHg) AVE (SD) | 144 (19) (n=39) | 124 (19) (n=35) | p=.001^* |
| Diastolic (mmHg) AVE (SD) | 77 (12) | 70 (12) | p=.003^* |
| pBP (mmHg) AVE (SD) | 68 (16) | 54 (16) | p=.0006^* |
| CRP (mg/mL) < 48H | 18 +/- 7.3 (6 tested) | 5.6 +/- 5.5 (12 tested) | p=.003^* |

**Table 3.** Relative risks by sex/race and conditions.

|  | RR severe outcome | P | 95% CI |
| --- | --- | --- | --- |
| Female | 0.8 | 0.5 | 0.55 to 1.32 |
| Black | 0.9 | 0.66 | 0.56 to 1.45 |
| Obesity | 2.04 | 0.0075 | 1.21 to 3.44 |
| T2DM | 1.29 | 0.1288 | 0.93 to 1.82 |
| HTN | 1.92 | 0.1390 | 0.81 to 4.59 |
| SBP ≥ 140 mmHg | 2.25 | 0.0005 | 1.42 to 3.55 |
| pBP ≥ 60 mmHg | 2.4 | 0.0029 | 1.34 to 4.19 |
| ACEi | 1.1 | 0.52 | 0.75 to 1.74 |

Pre-COVID essential HTN is reported in 83% patients, and proportions by race and sex are identical. Patients with HTN are older (average age 68 vs. 55 years) (p= 0.006, z=2.746) while similar median BMI (33 vs 31 kg/m^2^) are observed between groups with and without HTN (**Table 4**). The proportion of patients with HTN parallels outcomes severity (100% in fatal, 84% in non-fatal severe, and 76% in non-severe), without reaching statistical significance between severe vs non-severe (90% vs. 76%) (**Table 2**), and HTN is not a significant relative risk of severe outcomes, (RR 1.9x, 95% CI 0.8-4.6 p= 0.14) (**Table 3**). 82% of patients with HTN had pre-COVID anti-HT drugs, 43% in monotherapy and 39% in polytherapy and similar number and class of anti-HT drugs prescribed are found by race/sex (except a trend for more polytherapy in White males) (**Table 4**), and by outcome severity (**Table 2**, **Fig. 2**). No statistically significant difference of outcomes can be found by race/sex, age, conditions such as T2DM, CKD or by comorbidity indices, Charlson and Elixhauser (**Table 2**).

**Figure 2.**
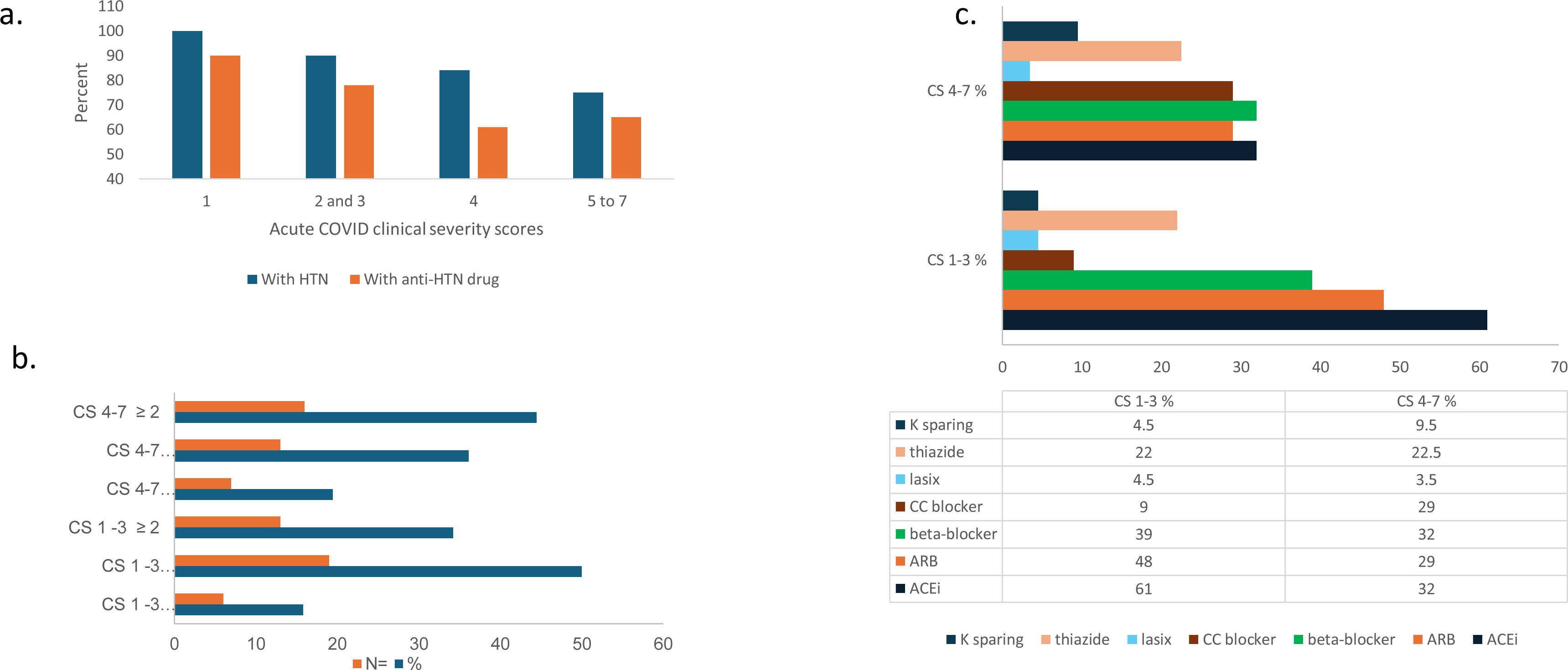
Comparison between ultimate COVID outcome and presence of pre-COVID HTN and HTN drugs prescribed, by number and class of drugs. a. Percentage of patients with pre-COVID HTN and anti-HTN therapy is proportional to COVID severity outcome. 1-death, 2-mechanical ventilation, 3-hi-flow oxygen, 4-low-flow oxygen, 5-admitted, did not require oxygen, required medical care, 6-admitted for non-medical reasons, 7-ED visit 24-48hours (n= 3). Note y axis starts at 40. **b. Number and proportion of subjects with pre-COVID HTN / anti-HTN drug prescribed, grouped by ultimate COVID outcome.** More subjects who experienced severe COVID were prescribed pre-COVID monotherapy for HTN, while subjects with non-severe COVID had anti-HTN multitherapy. **c. Antihypertensives prescribed to ClinSeqSer subjects prior to COVID, grouped by drug class and ultimate COVID severity.** Anti-HTN drugs prescribed to ClinSeqSer subjects pre-COVID grouped by action (vertical axis), and clinical score. We observe an enrichment of ACEi use in the group of subjects who experienced severe COVID outcomes but not a statistically significant difference. Proportions presented here represent [all drugs prescribed / all subjects with pre-COVID HTN, with or without anti-HTN therapy]. ACEi= angiotensin converting enzyme inhibitor, ARB= angiotensin receptor antagonists.

**Table 4.** Distribution of pre-COVID HTN and, if HTN, treatment prescribed, for overall cohort then grouped by race, sex. a. Proportion of patients with HTN by race, sex, age and BMI. b. Anti-hypertensive management of patients by anti-hypertensive drugs number, race and sex. HTN=hypertension, F=female, M=male. Anti-hypertensive=anti-HT.

|  | HTN | No HTN | p |
| --- | --- | --- | --- |
| **Black + Lax patients** | 84% | 16% | ns (B vs W) |
| **White patients** | 73% | 27% |  |
| **Sex Female/male** | 78% / 73% | 22%/27% | ns |
| **Age (years)** | 68 | 55 | 0.006 |
| **BMI (kg/m<sup>2</sup>)** | 33 | 31 | ns |

|  | anti-HT drug |  |  |
| --- | --- | --- | --- |
|  | no | one | ≥ 2 |
| **All %** | 18 | 43 | 39 |
| **B F %** | 22 | 41 | 37 |
| **B M %** | 10 | 53 | 37 |
| **W F %** | 25 | 37.5 | 37.5 |
| **W M %** | 19 | 62 | 0 |

Admit BP analysis is performed on 74 patients, 38 severe, and 36 non-severe, including 34 and 30 with prior COVID HTN respectively. Admit SBP average values are significantly higher (p= 0.001) in subjects with severe vs non-severe outcomes (144 vs 124mmHg) (**Table 2 and Fig. 3a**). The proportion of patients with admit SBP ≥140mmHg is higher in severe (∼60%) vs non-severe (∼19%) outcomes (Fig. 3c), whereas the proportion with lower admit SBP (100 to 129mmHg) is higher in non-severe (43%) vs severe (20%) (**Fig. 3c**). By ROC analysis, admit SBP ≥140mmHg correlates with severe outcomes with Se=75%, Spe=66%, and AUC (accuracy)=66% (**Fig. 4a**). Patients with admit SBP ≥140mmHg have a 2.25 relative risk of severe outcome (p=0.0005; 95% CI 1.42-3.54) (**Table 3**), and reach a severe clinical state earlier (within 5 days of admission, 80% vs 50% with admit SBP ≥ and <140mmHg respectively (Kaplan Meier graph / log-rank test p= 0.0093) (**Fig. 4b**).

**Figure 3.**
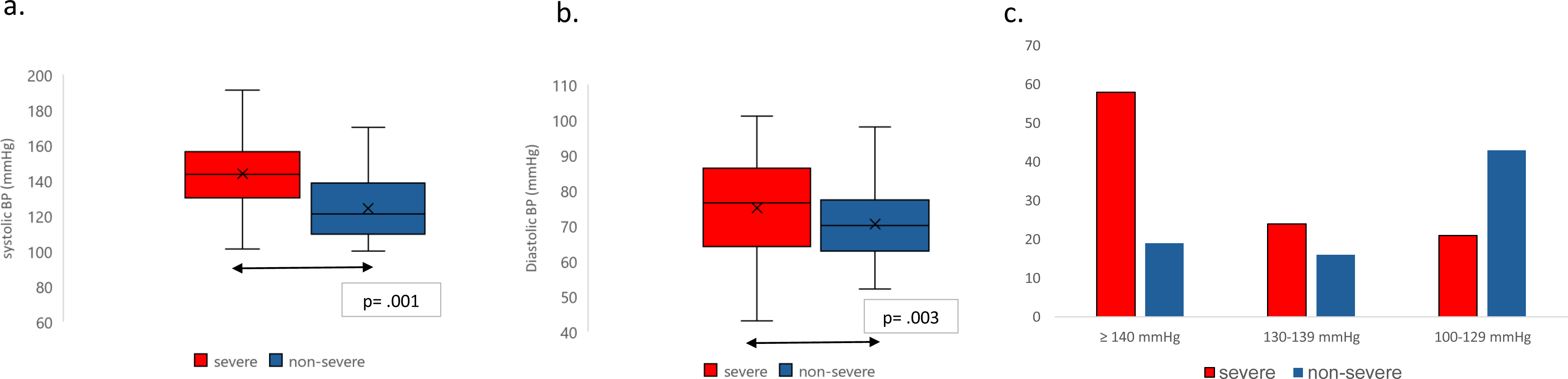
Admit systolic (a) and diastolic (b) blood pressure medians versus ultimate COVID severity. SBP median: 144 mmHg in fatal & severe (red) vs 118 mmHg in non-severe (green) COVID. vertical axis = median systolic SBP (a) or DBP (b), horizontal axis = COVID severity group. SBP= systolic blood pressure, DBP = diastolic blood pressure. P values calculated using Mann Whitney U test. **(c) Distribution of admit systolic blood pressure (SBP) values in each COVID severity group (severe vs. non-severe)**. Percentage of subjects with high, medium, and low admit SBP, grouped by ultimate COVID severity (vertical axis= patients %).

**Figure 4.**
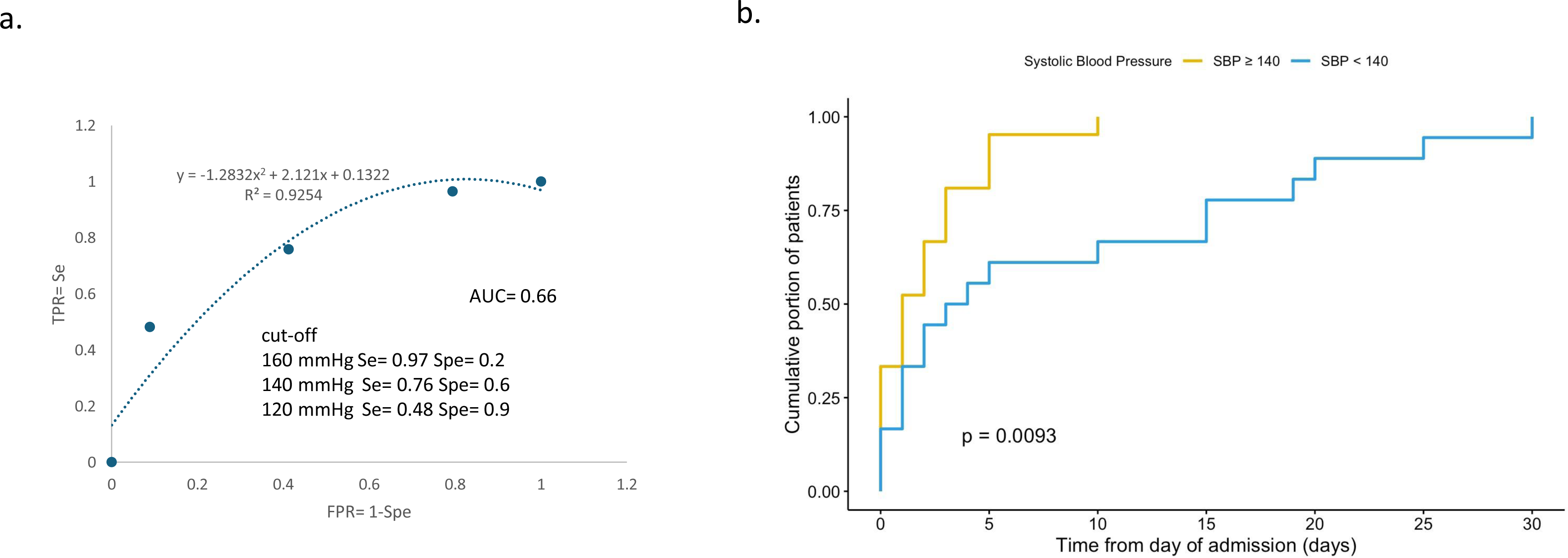
a. ROC curve graph of SBP threshold and AUC. b. Kaplan-Meier curves. Illustrating (vertical axis) cumulative number (proportion) of patients reaching a state of clinical severity (fatal and severe) from admission over (horizontal axis) days 0 to 50 of admission. Patients with admit SBP ≥ 140 mmHg (yellow line) and < 140 mmHg (blue line) with statistical significance by log rank test.

DBP median is 76 vs 68 mmHg in severe vs. non-severe (p= 0.03) (**Fig. 3b**). Pulse blood pressure (PBP= SBP minus DBP) is higher, 68 vs. 54 mmHg in severe vs in non-severe patients (p=0.0006), with significant difference of proportions of patients with PBP ≥ 60mmHg with severe (vs non-severe) clinical score (z= 3.42; p= 0.00062) with a 2.4x relative risk of severe outcome (p= 0.003; 95% CI 1.34-4.2) (**Table 2 and 3**). Of note, admit SBP did not correlate with prior HTN, number of anti-HT drugs (**Fig. Supp. S1**) although a trend toward more monotherapies could be observed with higher SBP on admission in patients with severe outcomes (**Fig. Supp. S2**).

Onset symptoms, analyzed by clusters, Hgb oxygen saturation (pulse oximetry) and oxygen supplement required over the first 24 hours of admission, did not correlate with ultimate COVID outcome (**Fig. S3 and S4**). First 24h oxygen requirements for 27 of 42 subjects with severe COVID outcome were indistinguishable from subjects with non-severe COVID outcome, indicating that 83% of cohort subjects appeared “non-severe” over at least the first day of admission. (**Fig. S5**). Only CRP values over admission correlated with outcomes (**Supp. Fig. S6**). However, CRP, eGFR (**Fig. S7**) or BNP (**Fig. S8**), were not helpful as early predictors [Refer als to Supp. File #2 bioMarkers].

## DISCUSSION

In this pre-vaccine COVID cohort, the prevalence of pre-COVID HTN is roughly twice that in LA residents [12]. In other early LA COVID data, by comparison, a rate of 60% HTN is reported among fatal COVID cases, out of 21,000 total cases [24]. Only an early study in Italy reported a similarly high rate of HTN (73%) in COVID patients with fatal outcome (average age 79 yo) [25] [26]. In an early COVID cohort in Wuhan, China, by Caillon et al, [27] much lower pre-COVID HTN rates are observed (28% among non-fatal, 59.5% among fatal, 36% overall), while higher admit SBP correlated with fatal outcomes vs. discharge (137 vs. 125 mmHg). The Ochsner health system, a neighboring uptown NOLA private Health system, reported that patients admitted for acute COVID had a much lower HTN prevalence, 34% in Black patients and 24% in White, and similar admit SBP (133mmHg). Notably, both cohorts have the same Urban NOLA demographics, proportions of obese patients and low CCI scores (<5) [28]. Since our cohort’s demographics reflect exactly that of the population of greater urban NOLA it is likely that the very high HTN prevalence in our cohort reflects higher social vulnerability of patients recruited in the downtown health systems (30% Black patients on Medicaid) compared to uptown (10% Black patients on Medicaid). It is known that HTN prevalence parallels higher socio-economic vulnerability [29]. Consistently, in a Black cohort recruited from NOLA churches, a prevalence of 73% hypertension is reported [30].

This study’s weakness is inherent to the small numbers of patients that may have led to skewed proportion of patients with HTN and of BMI values, obscuring correlation between HTN, anti-HT management and obesity. The trend of lower prevalence of obesity and HTN, respectively, in non-severe versus severe suggests that obesity’s contribution to COVID severity may be more important. Obesity, with 2.2 risk factor of severe outcome and a fair correlate of outcome with 70% accuracy by ROC analysis. This also may be related to the cohort’s anti-HT treatment, where correlation becomes less evident (**Fig. 3**)[31]. However, under-management of pre-COVID co-morbidity, including HTN is suggested by the high prevalence of HTN and a trend towards higher proportion of anti-HT monotherapy in patients with severe outcomes. This is likely amplified by poor (anti-HT) treatment compliance, well recognized in LA, and a correlate of high social vulnerability [32] although the classic additional secondary causes of high SBP (tobacco, alcohol, salt consumption, hematocrit, and steroids) were not explored. On the flip side, better pre-COVID HTN management, the trend observed of higher proportion of multiple anti-HT drugs in patients with non-severe outcomes, may have contributed to better COVID outcomes, overriding high co-morbidities impact as seen for nearly half of patients in our cohort [33]. As reported by others in HTN related death [29] we observed “stronger, unmeasured determinants of HTN and life expectancy” that may explain the surprisingly similar proportions of co-morbidities in Black female patients with severe or non-severe outcomes, such as polypharmacy, including drugs with respiratory depressant side effects and frequent prior total abdominal surgeries, (which can generate abdominal structural defects causing ineffective tachypnea and cough). Prior studies have raised similar concerns in similar population [9, 34].

Strengths of the study include focused time/region under-represented vulnerable population reflected by high obesity and HTN, in-person recruitment and comprehensive data collection by the same health professionals involved in patients ‘care at the same time. Another interesting observation from our study is the overlap of onset symptom cluster and early admit oxygen requirements between subjects who will experience severe versus non-severe outcome. This is a rarely reported event, possibly noticed only because of the sequence and depth of data collected, with most studies if overall admission outcomes parallel admit severity. This highlights the challenges inherent in predicting COVID clinical course early during admission. We deliberately opted for study of severe versus non-severe outcomes, rather than death versus discharge which does not discriminate patients by high vs. low level of care requirement.

In this cohort, common correlates of outcomes, such as HTN, T2DM, co-morbidity indices, clinical score systems, oxygen requirement or hemoglobin oxygen saturation were not significantly predictive of outcomes. While the CDC and the NIH recognize and support the studying special populations [35], the report of data originated from small(er) dataset triggers bluntly dismissive peer reviews. While the belief that large populations and dataset guarantee strong statistical significance is challenged, “the massive size of such databases is often mistaken as a guarantee for valid inferences” [36], peer review of data generated from small cohorts’ argue of “statistical weakness” that leads to blunt rejection. Rare publications are focused upon alternate methods to apply for studying small populations [37]. Emphasizing that this cohort was constituted at the very beginning of the pandemic with in-person recruitment in full PPE and the ultimate witness of times where 17% of patients admitted had fatal outcomes does not override the statistical strength dogma. We refer to the Bogalusa Heart Study and another study performed in Black community from NOLA [38] to highlight the context of major comorbidities experienced by the Black population in the State. Our snapshot of health documents that social vulnerability experienced by Black patients likely explains why similar co-morbidity can be observed a decade earlier than in White people. We therefore aimed to find a clinical predictor better adapted to this under-represented population with high social vulnerability, highly enriched for pre-COVID HTN and in the pre-vaccine period of the pandemic. Se, Spe and accuracy are low by ROC analysis, likely due to small numbers, but ROC prediction is also not suitable for clinical criteria and depends upon circumstances of implementation [39]. Because nearly 70% of patients with severe outcomes present with a non-severe respiratory state, admit SBP could be a simple yet valuable correlate to help triage patients in critically challenged healthcare systems.

The strength of the study is first to highlight that essential HTN does not always constitute a risk of severe outcome, possibly related to prior anti-HT drug management, and second, that higher SBP is correlated with more severe acute COVID outcome. In a early pandemic study in Wuhan, China, Caillon et al. report higher sBP on admission in acute COVID patients as a correlate of discharge [27]. In our cohort, we found in addition that admit SBP ≥140 mmHg in association with higher and earlier relative risk of severe outcome that may constitute a simple, early, and low-cost clinical correlates of severity for COVID, and worth exploring in other acute viral illnesses.

The correlation between high admit SBP with severe outcomes corroborates the findings by Park et al. [40] of abnormal left ventricular global longitudinal strain predicted worse outcomes in patients hospitalized with COVID as its impact upon sBP was found associated with stage I hypertension [41]. In COVID the “degradation of lung function that could associate with rise in BP” as suggested by Vicenzi et al. [42]. A pulse blood pressure over 60mmHg is a risk factor for heart disease, and correlates with stiffness of the body’s largest arteries in older adults [43]. Other hypotheses and further studies regarding why high admit SBP is associated with worse COVID outcome could be explored including: 1) Pre-COVID comorbidities, such as poorly controlled HTN, CKD, obesity, polypharmacy including ACEi, advanced age, or, 2) a specific effect of SARS CoV-2 on the renin angiotensin axis, such as direct infection/kidney injury (reflected by accumulation of BNP) [42], or, 3) specific polymorphisms, frequently encountered in Black populations, of sodium channel (Liddle phenotype), or hypersensitivity to bradykinin (ACEi induced angioedema) [44, 45] or specific variants of the ACE2 receptor [46] or, 4) Specific strain of SARS CoV-2 spreading at that time responsible for increased admit SBP in a subset of subjects. One also could hypothesize that high admit SBP may be a risk for developing long-COVID [47] and contributing to recently reported damage to the blood brain barrier [48].

## 3. Conclusions

Overall, we conclude that, among patients admitted early in the pandemic, enrichment with pre-COVID HTN (83%) reflects the population’s very high social vulnerability. Obesity, admit BP and CRP correlated with severe outcome whereas age, pre-COVID HTN, anti-HT, T2DM, and admit oxygen saturation did not. Our study shows that increased admit SBP is independent of pre-COVID HTN and anti-HT drugs. This small dataset may serve as a useful indicator to regional guidelines anticipate similar outbreaks in the future and raise awareness of possible worsening of regional HTN in underrepresented populations.

## Supporting information

Suppl. results bioMarkers

## Data Availability

The presented data will be shared on reasonable request to the corresponding author.

