## Supplementary material for "Admission systolic blood pressure and obesity correlate with fatal and severe acute COVID-19 in the population of New Orleans, LA": Suppl Tables and Figures

Supp. Tables and Figures

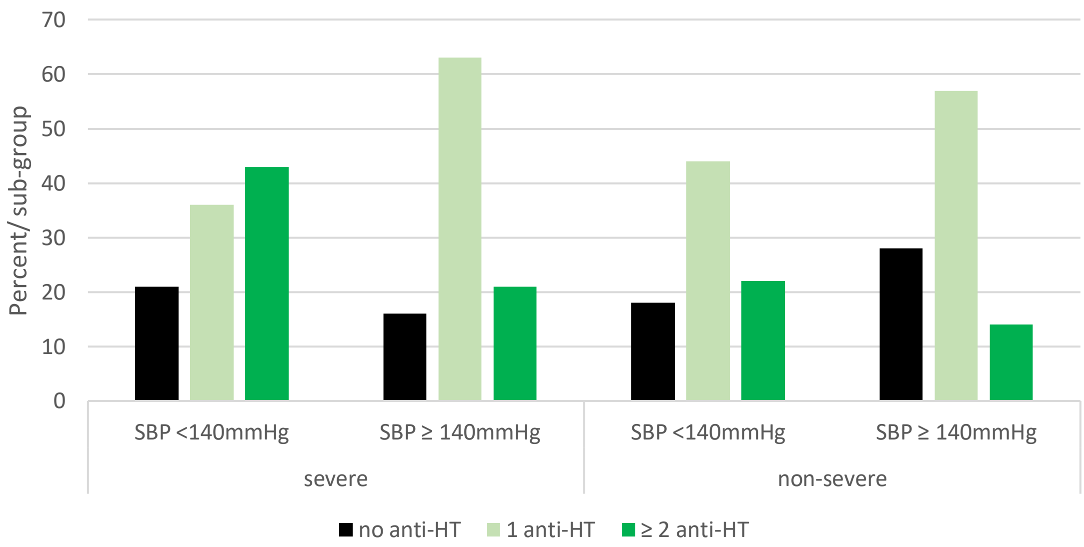


**Suppl. Fig. 1** Repartition of anti-HT drugs management by outcomes and by admission SBP levels.

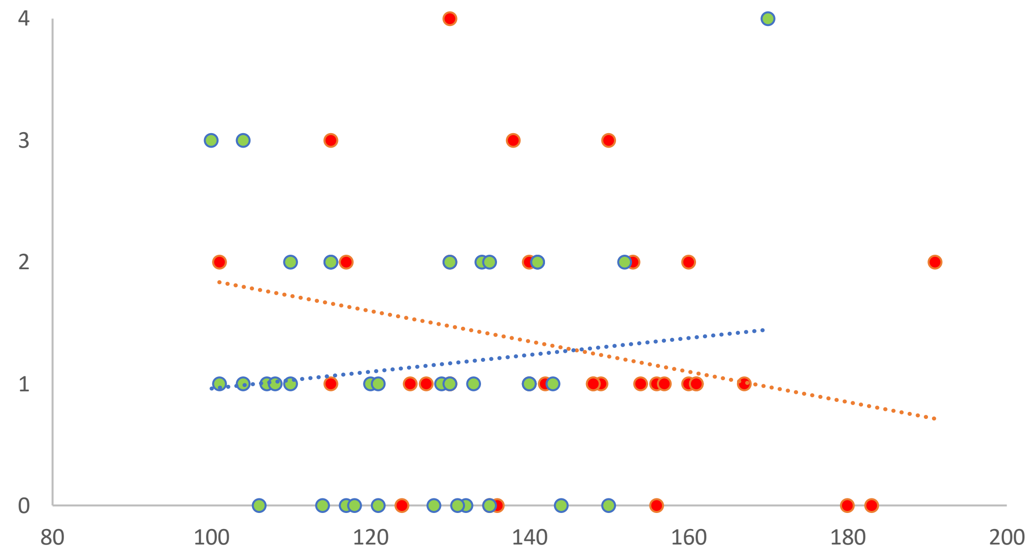


**Supp. Fig. 2** Number of anti-HT drugs vs. SBP values on admission in patients with severe (red) and non-severe (light green) outcomes.

| Onset of symptoms cluster | Flu like + SOB (core) | core + fatigue + AMS | productive cough | gastro-intestinal | pauci-symptomatic | afebrile | confusion |
|---|---|---|---|---|---|---|---|
| Percentage | 19% | 21% | 9.5% | 26% | 0.2% | 17.5% | 6% |

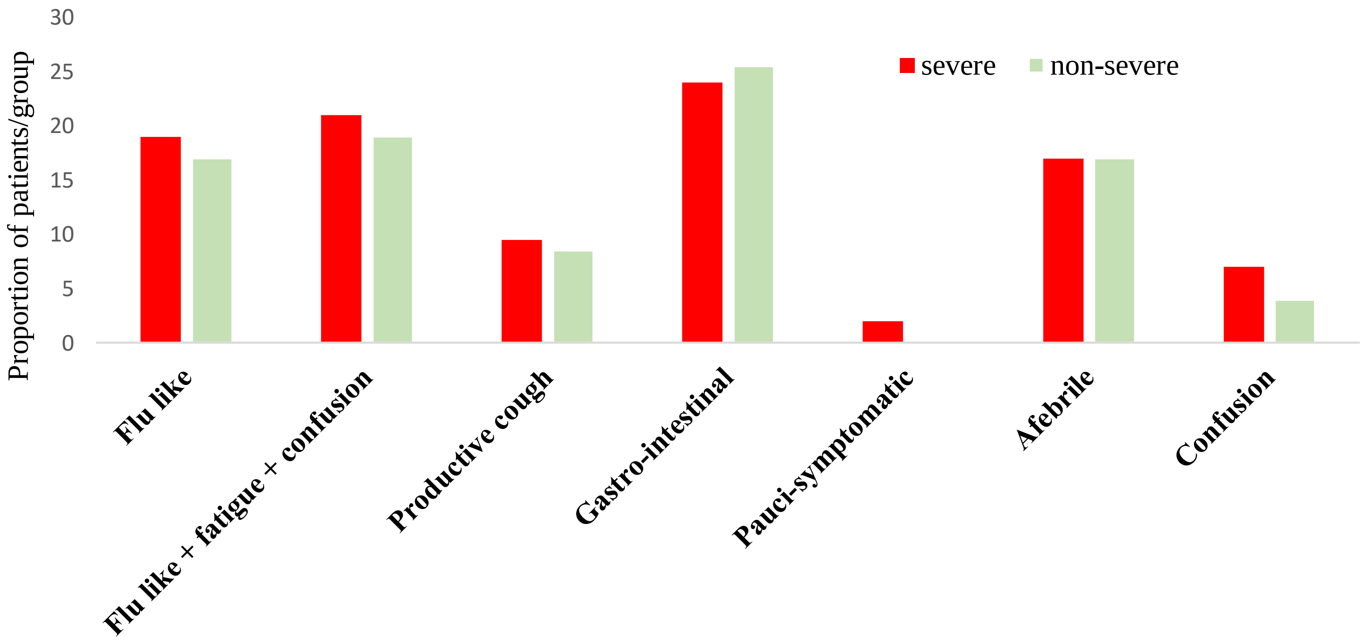


**Supp. Fig. 3. Symptoms over onset period.** a. Proportions of patient in each clinical cluster of symptom onset.
b. Repartition of clusters by ultimate COVID outcome. Horizontal axis= onset symptom clusters, vertical axis= percent of subjects complaining of that symptom cluster on initial presentation. Patients with severe (red histogram) and non-severe (green histograms). SOB: Shortness of Breath.

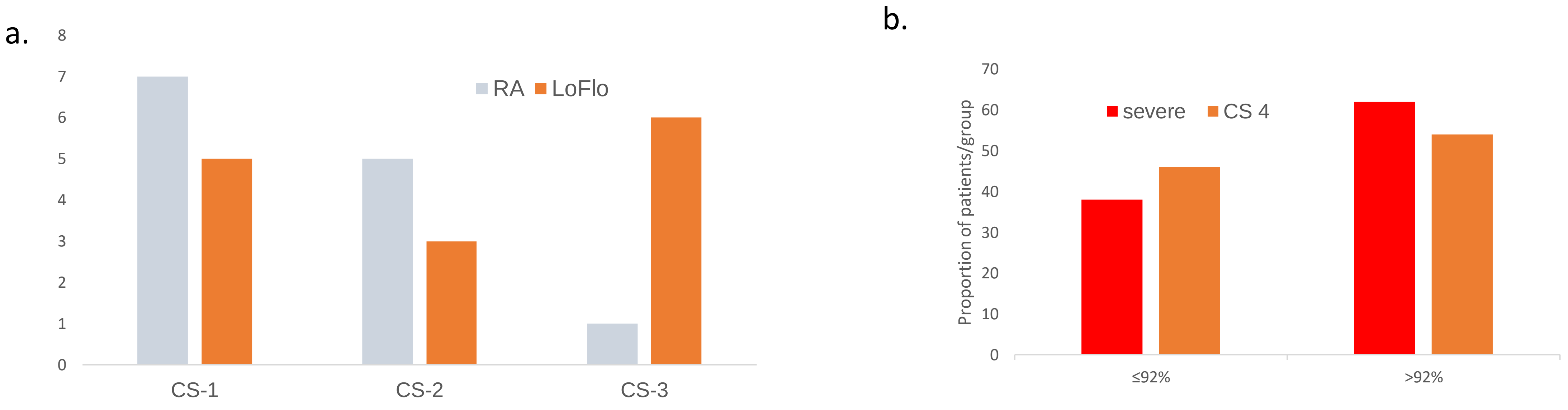


**Supp. Fig. 4. a. Early oxygen requirement (first 24 hours) of admission does not predict the ultimate severe (CS 1-3) outcome of patients. Vertical axis**: number of subjects, **horizontal axis: initial** COVID clinical severity score grouped by initial oxygen requirement (RA= room air, LoFlo <6L/min) in patients with ultimate outcome: CS-1: fatal, CS-2: required mechanical ventilation, and CS-3: required HiFlo oxygen. **b**. **On-admit oxygen saturation compared to ultimate COVID outcome.** Acute COVID outcome according to oxygen saturation on admission (reportable* SaO2 of 34 patients /42 in severe (CS 1-3) and 26/26 non-severe (CS 4). Patients were classified in groups according to their oxygen saturation (SaO2 ≤92% and >92%) on admission Ultimate severe: red histograms and Low flow oxygen (CS-4): green histograms.

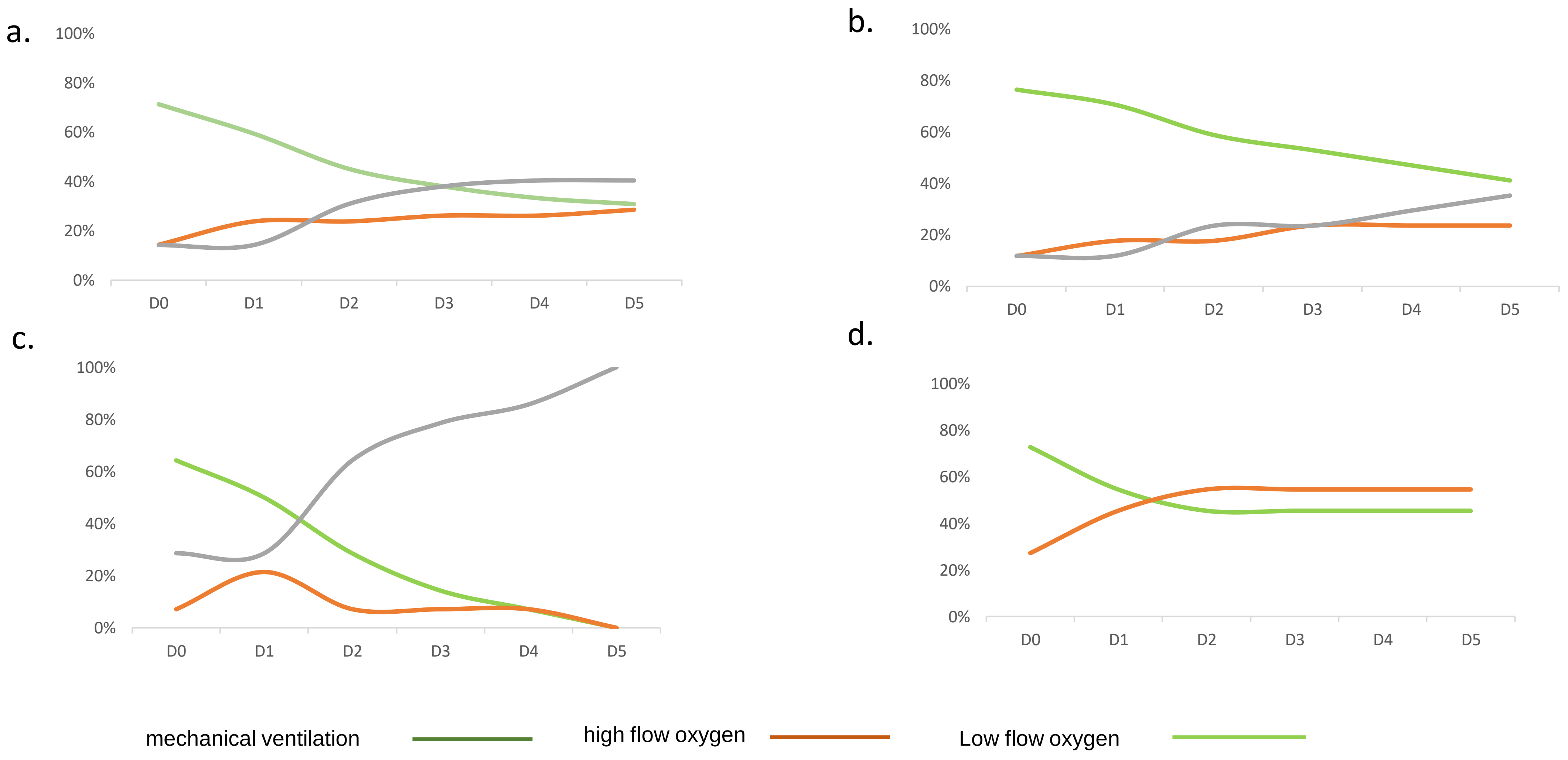


**Supp. Fig. 5. Proportion of patient with ultimate severe outcomes (vertical axis) daily clinical evolution (horizontal axis: Day 0=admission to Day 5) by oxygen requirement (lines color: dark green patients requiring mechanical ventilation, light green patients with high flow oxygen and orange, patients with low-flow oxygen). Group of patients by ultimate outcome: a.** severe outcome; **b.** fatal outcome ; **c.** mechanical ventilation / non-fatal; **d.** high flow oxygen. The curves illustrate that the majority of patients with ultimate severe outcomes experienced non-severe outcome for the first days of admission. This is particularly striking in the group of patients with fatal outcomes (7b.)

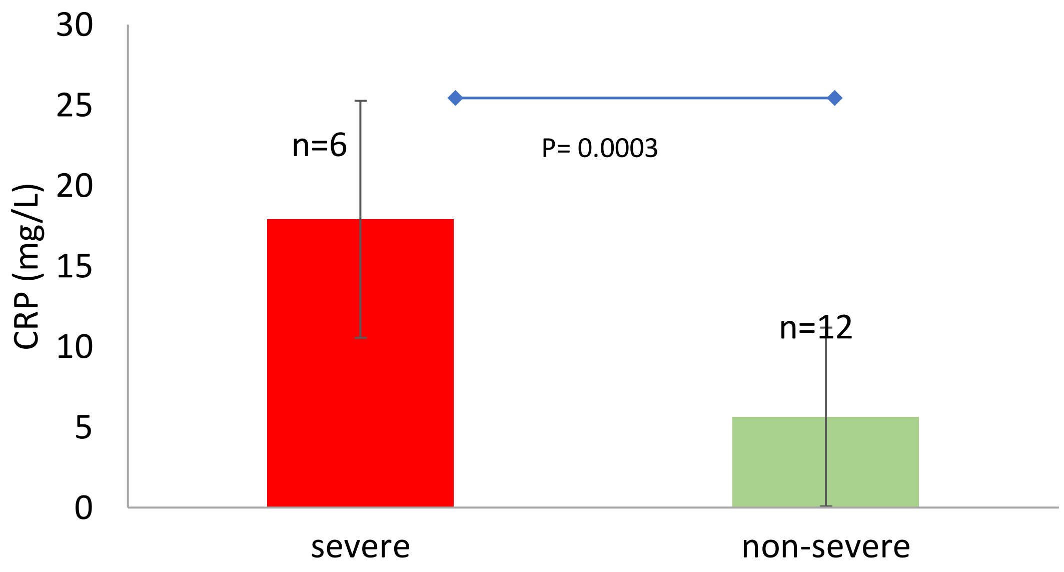


**Supp. Fig. 6. CRP (adult ref. range < 10 mg/L) values collected over the first 48 h of admission for ClinSeqSer acute COVID subjects.** A subset of subjects had CRP collected during first 48h of admission 6/42 severe, v 12/47 non severe. Vertical axis= CRP values, horizontal axis= clinical severity. Bars=SD, p value: students t test, 2 tails, unequal variance.

a.

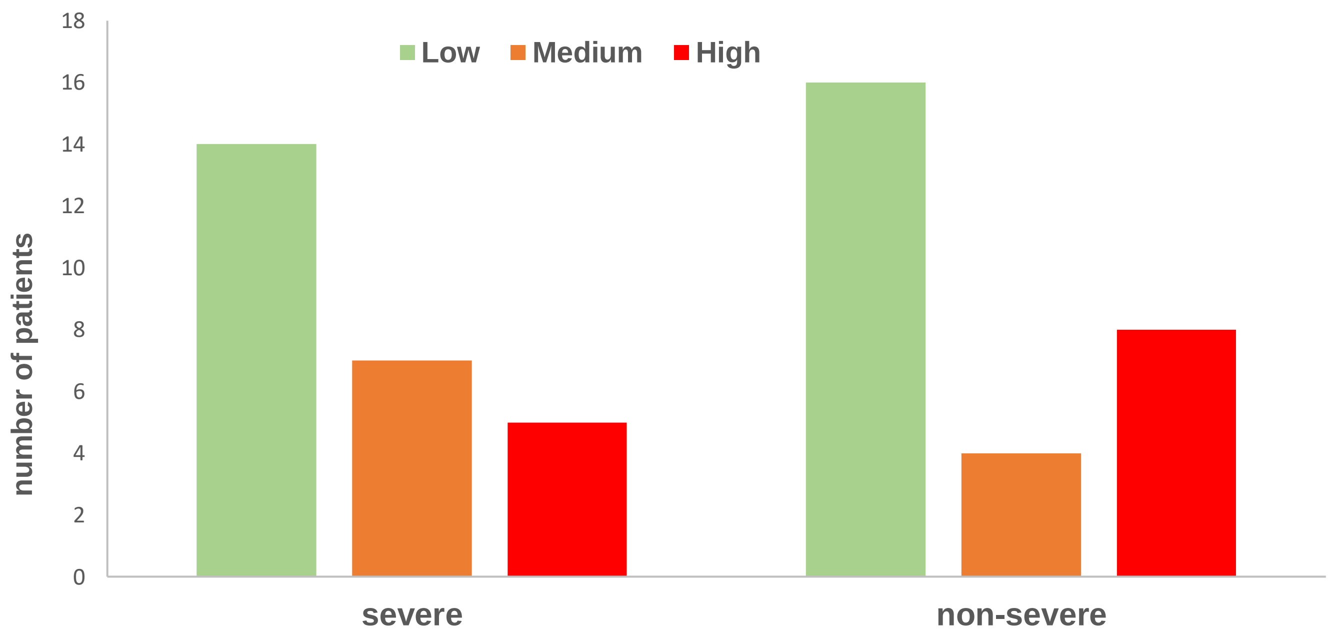


**Supp. Fig. 7. Heart failure likelihood scored by age and BNP worst value over admission versus ultimate COVID outcome of severe versus non severe.** Likelihood range after the Heart failure likelihood score by age and BNP values, after the guidelines of the Heart Failure Society / European Society of Cardiology (Ref Methods Suppl Table XXX). Heart failure likelihood histograms color coded as green for low, orange for medium and red for high. Horizontal axis= acute COVID severity group. Vertical axis= number of patients tested over admission.

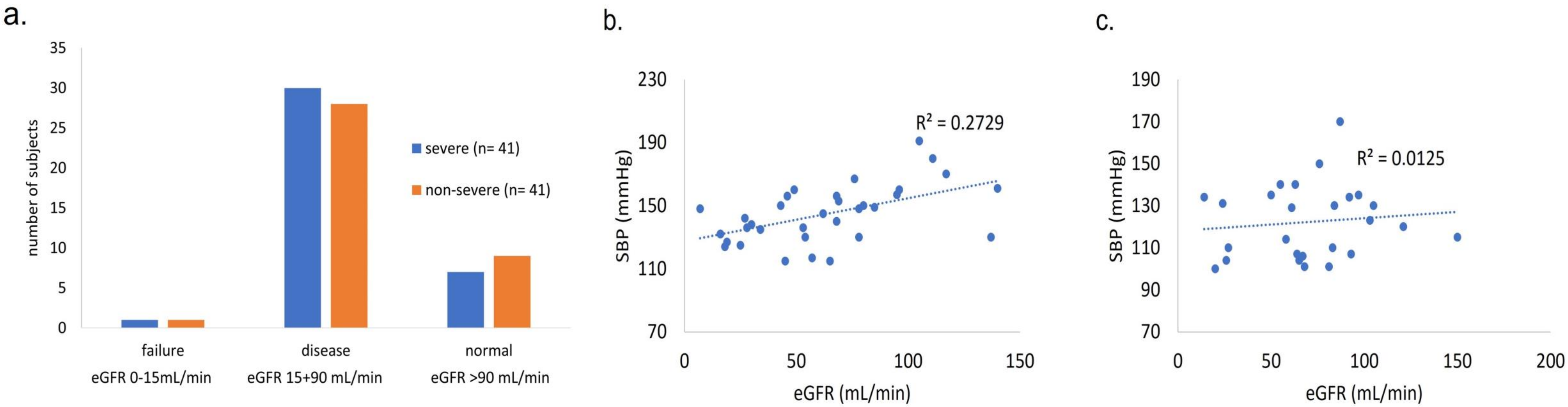


**Supp. Fig. 8. Admit Kidney Function did not predict acute COVID severity**. a. Kidney function/eGFR levels on admission for acute COVID.. b eGFR versus SBP among subjects with severe COVID outcome. c. eGFR versus SBP among subjects with non-severe COVID outcomes.
