## Supplementary material for "Admission systolic blood pressure and obesity correlate with fatal and severe acute COVID-19 in the population of New Orleans, LA": Suppl. Methods

Supplemental File 1 Methods, Definitions, and methodology.

Study type. ClinSeqSer is an observational, longitudinal, regional study, initiated March 2020, aimed to define correlates of acute COVID outcomes with any of the following: pre-COVID Long Term Conditions (LTC) and medications, presenting vitals, clinical evolution during admission, and clinical laboratory results. Recruitment: Adult patients ≥18 years old were approached based upon positive nasopharyngeal SARS CoV-2 PCR, followed by informed consent. Enrollment occurred in person during hospital admission.

Ethics approval. Study was conducted with approval from Tulane University School of Medicine Institutional Review Board (IRB) under protocol “Collection of SARS CoV-2 Serum and Secretions for Countermeasure Development” [IRB 2020-396, approved 3/20/2020]. All procedures were performed in accordance with ethical standards of Tulane IRB and Helsinki Declaration of 1975 (revised 1983).

Clinical trials registration number: NCT04956445.

Definitions and methodology, including the Clinical Scores stratification, per WHO clinical score evaluation on a seven-point ordinal scale (see Supplemental Information). **Data source and collection.** Enrollment sites included Tulane Medical Center (TMC) and University Medical Center (UMC), both in NOLA. Clinical data and laboratory test results were collected from electronic health records (EHR) MEDITECH (TMC) and EPIC (UMC) and stored in REDCap. To optimize homogeneity and accuracy of data, extraction was performed with entry verification for ~20% of cases, randomly selected, verified by independent study team members, and correlation debriefing meetings performed regularly.

**Statistical analysis:** We report categorical variables as percentages, continuous variables as mean/SD if normally distributed or median/IQR if non-normally distributed. Data were analyzed using Chi-squared, Mann Whitney U test or t-test methods as appropriate. Statistics applied for normal distribution assessment, average/median/IQR, ROC curve analysis, regression analyses were performed in Excel and: Statology for t-test, Mann Whitney, two proportion Z-test calculator, Statistics Kingdom for Kaplan-Meier log-rank test [20-22]. For relative risk we used the calculator from MedCalc [https://www.medcalc.org/calc/relative_risk.php].

State Health Score [1]: Score is based on 21 metrics, extracted from datasets of the Center for Disease Control (CDC), Kaiser Family Foundation (KFF) and Substance Abuse and Mental Health Services (SAMSHA), and including disease risk factors and prevalence, substance abuse, lifestyle habits and health outlook.

Wealth and Social Vulnerability Index (SVI) were compared between racial/ethnic groups using ZIP codes and health care coverage status collected on recruitment. Socio-economic status is estimated by first five digits of ZIP code, which reflects habitat’s ground levels / flooding risk by Federal Emergency Management Agency (FEMA) maps and proportion of inhabitance by Black/White residents by US census. SVI scores are a nationwide estimate by region / ZIP code [2, 3].

ZIP code, Insurance, Race/sex were collected from EHR. Because of age and comorbidity similarities, we included the two Latinx patients within the group of Black patients.

Body Mass Index (BMI). Collected from admission or medical visits within the six months preceding COVID admission, kilogram weight/square meter height (kg/m^2^). Obesity is defined by WHO criteria as BMI ≥30kg/m^2^, Super-obese as BMI ≥40kg/m^2^.

**Essential Hypertension (HTN).** Defined as systolic blood pressure (SBP) ≥130 mmHg (2018 American College of Cardiology (ACC) guidelines [4]), with ICD10 code for HTN, present prior to COVID diagnosis, reported in pre-COVID clinical notes in two prior visits. Medications prescribed for treatment of HTN were also collected from EHR. For patients without prior visits recorded, presence of HTN/anti-HT medications was collected from History and Physical (H&P) on admission and if available, from prior admission H&P notes, as in a disease management program.

Co-morbidities. Comorbidities included in indices calculation and elsewhere were diabetes, dyslipidemia, chronic kidney disease (CKD), cardiovascular (CV) disease, heart failure (HF), atrial fibrillation (aFib), autoimmune disease (AID), cancer, major past surgery, psychiatric conditions, alcohol consumption, cigarette smoking, polysubstance abuse, opiate use disorder. Comorbidities and prescribed medications were collected from EHR.

Co-morbidity scores: 1- The Charlson comorbidity Index score (CCI) in the modified Deyo version is independent of age and score 1-29. 2- The Elixhauser Index score, van Walraven algorithm, score provided either by range of likelihood for in-hospital death, -19 (less) to 89 (more), or by percent of possible points 0 to 82.4%. Calculators for both scores are available free online at orthotoolkit.com.

Blood pressure (BP) on admission. Systolic and diastolic BP were collected from first vitals registered by emergency medical services or in emergency department. To avoid false high readings when regular size arm cuffs are used in obese patients, TMC and UMC adhere to policies requiring use of large arm cuffs. We excluded values when patients were mechanically ventilated prior to transfer from an outside hospital or profoundly hypotensive on admission (SBP ~90 +/-2 mmHg, MAP <70 mmHg), all confirmed in EHR.

Table Methods supp. Table 1

| Score | Definition | Note |
| --- | --- | --- |
| 1 | death within 28 days  or  death during initial COVID admission | Death occurred during acute COVID admission with confirmed diagnosis of COVID made by nasopharyngeal PCR, with symptoms of acute respiratory illness or death attributed to any clinical illness compatible with COVID according to treating team. |
| 2 | mechanical ventilation | Some patients required mechanical ventilation for extra-respiratory manifestations of acute COVID-19 (eg, airway protection in setting of altered mental status). |
| 3 | non-invasive ventilation/high flow oxygen ≥ 6 L/min | The cut-off of high flow oxygen at 6 L/min was chosen as it is used in participating hospitals as a threshold for initiation of high-flow nasal oxygen; we considered this level of support or more severe disease to approximate a score of 5–8 using the WHO scale |
| 4 | low flow oxygen < 6 L/min |  |
| 5 | admitted/did not require oxygen supplementation but medical care |  |
| 6 | admitted did not require oxygen supplementation or medical care | “social admission” |
| 7 | “not admitted” | 3 patients had stay in the emergency department of ~ 48 hours due to results of nasal swab PCR issued initially by the State Lab. over the first couple of months with a turnaround time of 48 hours. |

Clinical scores stratified per WHO *Clinical score evaluation* on a seven-point ordinal scale 1- 7 [19]. Severe and non-severe groups: patients with clinical scores of 1 to 3 are included in “fatal & severe”, hereafter grouped together as “severe” unless otherwise specified, and 4 to 7 in “non -severe” COVID outcome groups.

**Biomarkers (BNP, CRP, eGFR):** For biological test results, worst value during COVID admission was collected. Based on lack of normal distribution for BMI and BNP, median/interquartile ranges were reported, and Wilcoxon rank-sum (Mann-Whitney) test used to compare outcome subgroups.

**NT-proBNP score of heart failure likelihood score:** Heart failure likelihood score by age and BNP ranges (ng/L) from Heart Failure Society guidelines, European Society of Cardiology.

| **BNP (ng/L)** | < 300 | 300-450 | 300-900 | 300-1800 | >450 | >900 | >1800 |
| --- | --- | --- | --- | --- | --- | --- | --- |
| **age (year)** | any | <50 | 50-75 | >75 | <50 | 50-75 | >75 |
| **HF likelihood** | LOW | MEDIUM | | | HIGH | | |

2.	*FEMA Hazards Map*. Available from: <https://hazards.fema.gov/nri/map>.

3.	CDC. *ATSDR Place and Health Map*. 2024; Available from: <https://www.atsdr.cdc.gov/placeandhealth/svi/interactive_map.html>.

4.	Whelton, P.K., et al., *2017 ACC/AHA/AAPA/ABC/ACPM/AGS/APhA/ASH/ASPC/NMA/PCNA Guideline for the Prevention, Detection, Evaluation, and Management of High Blood Pressure in Adults: A Report of the American College of Cardiology/American Heart Association Task Force on Clinical Practice Guidelines.* J Am Coll Cardiol, 2018. **71**(19): p. e127-e248.
