## Supplementary material for "Admission systolic blood pressure and obesity correlate with fatal and severe acute COVID-19 in the population of New Orleans, LA": Suppl. results bioMarkers

**Suppl. File # 2 Bio Markers**

**BioMarkers and acute COVID Severity**

**NT pro-BNP**

Natriuretic peptide tests, NT-proBNP (pg/mL).

**Method:** Tests were run either on Roche, Elecsys or Abbott, Architect platforms. Both display close correlation, with similar upper range of 30,000 and 35,000pg/mL, with Roche allowing issue of results up to 70,000pg/mL after 2-fold dilution. For values markedly above the analytical upper range (provided by the manufacturer), to avoid skewing the median with outliers, we capped values at 35,000pg/mL. Since multiple studies have attributed a prognostic value to raw NT pro-BNP levels over COVID admission, we calculated the values of NT-proBNP median/IQR by group of severity. We pooled results by patients, not time from admission, because of NT-proBNP result availability, timing, and frequency of measures. NT proBNP values were available for 64% (56/89) of subjects, 30% (26/89) with severe and 34% (30/89) with non-severe outcomes. On day 1 of admission, NT-proBNP is measured for 4 severe and 4 non-severe subjects and for 11 subjects a first BNP is measured in the first 3 days of admission. Most of the subjects had NT-proBNP ordered after 4 days (up to 22 days) after admission, performed more than once and up to 4 times for 3 severe patients. For the scoring of heart failure likelihood in acutely dyspneic patients, among a plethora of interpretation guidelines, we took the worst value over admission and used the age-adjusted cut-off of NT-proBNP from the PRIDE and ICON-Reloaded Investigators [25].

**Results:**

**Likelihood of heart failure score in severe versus non-severe COVID outcome groups are presented (Fig. 9).** Correlation between age adjusted values of NT-proBNP versus likelihood of heart failure by severity is provided (Fig. S5b).No significant difference in number of patients with or without high likelihood of elevated BNP is observed in severe versus non-severe groups.

**Raw levels of NT pro-BNP:** In the group of subjects with severe outcome, the mean raw values of NT-proBNP measured day 1-3 is 9500pg/mL (range 0.5-35,000; median= 466; IQR= 16600) and signifcanlty higher than in non-severe value, 1800pg/ml (range 6-15,500; median=159; IQR= 1834).

**Discussion:** By comparing mean/median IQR of raw NT-proBNP values in severe vs non severe of anytime NT-proBNP, we found significant difference between severe and non-severe groups. Using the PRIDE age adjusted algorithm, however, we do not explain why we have similar proportions of likelihood of HF in patients with severe and non-severe outcomes. We note limitations to interpretation of those observations: first, few subjects have NT-proBNP ordered on admission and few had consistent dyspnea leading to marker order. Second, age adjusted BNP cut-offs leave large zones of indecision since we were unable to interpret values measured on/after days into admission, since interpretation requires baseline heart function, evolution of renal function, and differs in the setting of valve dysfunction, a PE, acute coronary syndrome, arrythmia, or volume overload, and with fluid balance, oxygen support and use of cardio-pressors. Multiple studies have reported NT-proBNP levels, independently of HF diagnostic, as strong correlate of severity for acute COVID. Due to multiple origins of NT pro-BNP elevation, and to small numbers, we cannot consistently conclude either a correlation between COVID severity and differences of NT-proBNP levels observed. Overall, our study raises questions on how ordering and interpretation of NT-proBNP can be better standardized for diagnosis of severity and management of acute COVID and viral acute respiratory infections in general.

**Glomerular Filtration Rate (eGFR mL/min)**

**Method:** The eGFR estimate provided in clinical laboratory results is calculated from serum creatinine using the MDRD equation (2006). Interpretation of values: 0-15mL/min, kidney failure; 15-90mL/min, kidney disease; > 90mL/min, normal kidney function.

**Results:** As kidney disease is commonly associated with high blood pressure, we questioned whether higher SBP on admit correlated with worse kidney function in the severe COVID outcome group. The first eGFR measured was recorded and stratified by kidney function (failure: eGFR <15mL/min, disease: 15-90 mL/min, normal: > 90mL/min) (**Fig. 10**). No significant difference was observed in kidney function in severe vs. non-severe outcome (Fig. S6a). Consistently, no correlation is observed between SBP and eGFR for either severe or non-severe COVID outcome subjects (Fig. S6 b,c).

**C reactive protein (CRP mg/mL)**

**Methods:** CRP values results of local clinical laboratory, with normal values being either <0.3mg/mL or 10mg/L. Only CRP values from blood draw within the first 48 hours of admission were assessed. We only collected values of CRP ordered within 48 hours of admission (**Fig. 11**), as the evolution of CRP over the entire admission is beyond the scope of this study.

**Results:** Only a subset of subjects had a CRP measured during the first 48 hours, 6 of the 42 severe and 12 of the 47 non-severe. The mean values for severe vs. non-severe were 18 mg/mL vs. 5.5mg/mL therefore significantly different by t test, 2 tails unequal variance (p= .0034).

**Discussion:** By comparing the raw CRP values in severe vs non-severe patients, we found a statistically significant difference by z score (Table 2). However, the CRP was rarely ordered within the first 24-48 hours, although the small number of levels obtained in this timeframe seem to correlate with COVID outcomes.

**Conclusion:** In this small study, biomarkers BNP, eGFR, and CRP did not conclusively correlate with COVID outcome.
